# Development and validation of a near-comprehensive RxNorm valueset of opioid medications

**DOI:** 10.1101/2024.11.05.24316759

**Authors:** Michael Wasz, Prabhu RV Shankar, Elizabeth Sprouse, Lyndsey Kirchner, Matthew H. Garber, James Jones, Kenneth D. Mandl, Andy McMurry

## Abstract

**Objective:** Develop a near-comprehensive opioid medications valueset for population measures of opioid related treatments and outcomes. The opioid valueset should be free, open source, and conform to the RxNorm standard federally mandated in every US-certified electronic health record.

**Materials and Methods:** Cumulus opioid valueset was manually curated by the authors and expanded using computer assisted curation. Opioid classifier rules were developed to select opioid RxNorm concepts with known opioid receptor interactions, ingredients, keywords, and drug product formulations. Twelve publicly available valuesets were used to develop and validate the Cumulus opioid valueset. Validation accuracy was measured against a corpus of opioid medication orders and non-opioid pain relievers.

**Results:** Cumulus opioid valueset recall was >99.9% when measured against opioid prescription RxNorm codes from UC Davis Health and Brigham and Women’s Hospital. Cumulus opioid valueset was 100% specific compared to three valuesets of non-opioid pain relievers.

**Discussion and Conclusion:** To the authors’ knowledge, Cumulus opioid valueset is the largest publicly available valueset of opioid medications (8,926 RxNorm concepts). The intended use of this opioid valueset is for population health measures of opioid medications and related patient outcomes.

## INTRODUCTION

Opioid-based analgesia is the most commonly used treatment in postoperative pain management, with more than 95% of surgical patients receiving opioids during hospitalization[1,2]. This accounts to roughly 55 million patients in the United States each year. While many patients treated with opioids recover well, others experience opioid related adverse events[2] or develop opioid use disorder(OUD) [3,4]. To measure the frequency of opioid prescriptions[5,6] and opioid related outcomes[7], a comprehensive vocabulary[8] of opioid medication concepts are needed.

Electronic health records (EHR) contain longitudinal patient history[9–11], enabling measurements of patient status before, during, and after medication orders containing opioids, [12,13]. By analyzing the frequency of patient care trajectories involving opioid medications, it should be possible to decipher which care trajectories most commonly lead to better or worse outcomes. EHR data is a valuable resource to measure outcomes of opioid use disorder treatments such as naloxone and buprenorphine. At the time of this study, there are at least 30 opioid valuesets listed by the US national library of medicine (NLM)[8]. However, it remains unclear which combination of opioid valuesets should be selected to maximize sensitivity and specificity in opioid medication studies.

The primary objective of this study is to develop a near comprehensive list of opioid concepts referencing RxNorm[14]. RxNorm is the federally mandated standard drug vocabulary in the United States Core Data for Interoperability (USCDI)[11]. RxNorm contains precise drug type information, including brand names, generic names[15], medication strength, prescribable drug names[16], and relationships[17,18] to drugs across thirteen common medical vocabularies. The secondary objective of this study is to enable “push button population health” [9] measurement of opioid prescriptions and outcomes at national scale by adopting 21st Century Cures Act [19] mandated health data USCDI standards[11].

This study reports the Cumulus opioid valueset code[20] and data[21], freely available under the Apache-2.0 open source license. The Cumulus opioid valueset is published on VSAC[22].

## METHODS

The Cumulus opioid valueset was developed using published sources of opioids[23–30], expert manual curation, and computer assisted curation (**Figure 1**). Published valuesets were downloaded from the Value Set Authority Center (VSAC)[8]. Authors manually reviewed each opioid entry from an opioid medication valueset[29] using the Unified Medical Language System (UMLS) and Bioportal[31]. Each opioid concept was expanded to a larger valueset of opioid concepts having one or more opioid ingredients. Authors then manually curated a list of opioid ingredients, opioid keywords, and opioid classification rules for computer assisted curation. The medication reference terminology[32–34] (MED-RT) was used to automatically select all RxNorm concepts with known opioid receptor interactions[35,36]. Published opioid valuesets[23–30] were processed using opioid ingredients, opioid keywords, and opioid classification rules. The valueset aggregation of RxNorm concepts selected for opioid ingredients, opioid keywords, opioid receptor interactions, and opioid classifier rules were used to compile the Cumulus opioid valueset. Three VSAC non-opioid valuesets[37–39] were analyzed to provide examples of medications that do not include opioids. Non-opioid valuesets were used to test if non-opioid concepts were incorrectly included in the Cumulus opioid valueset. Lastly, sources of opioid prescriptions were analyzed to validate real world use of Cumulus opioid valueset in two medical centers.

**Figure 1:**
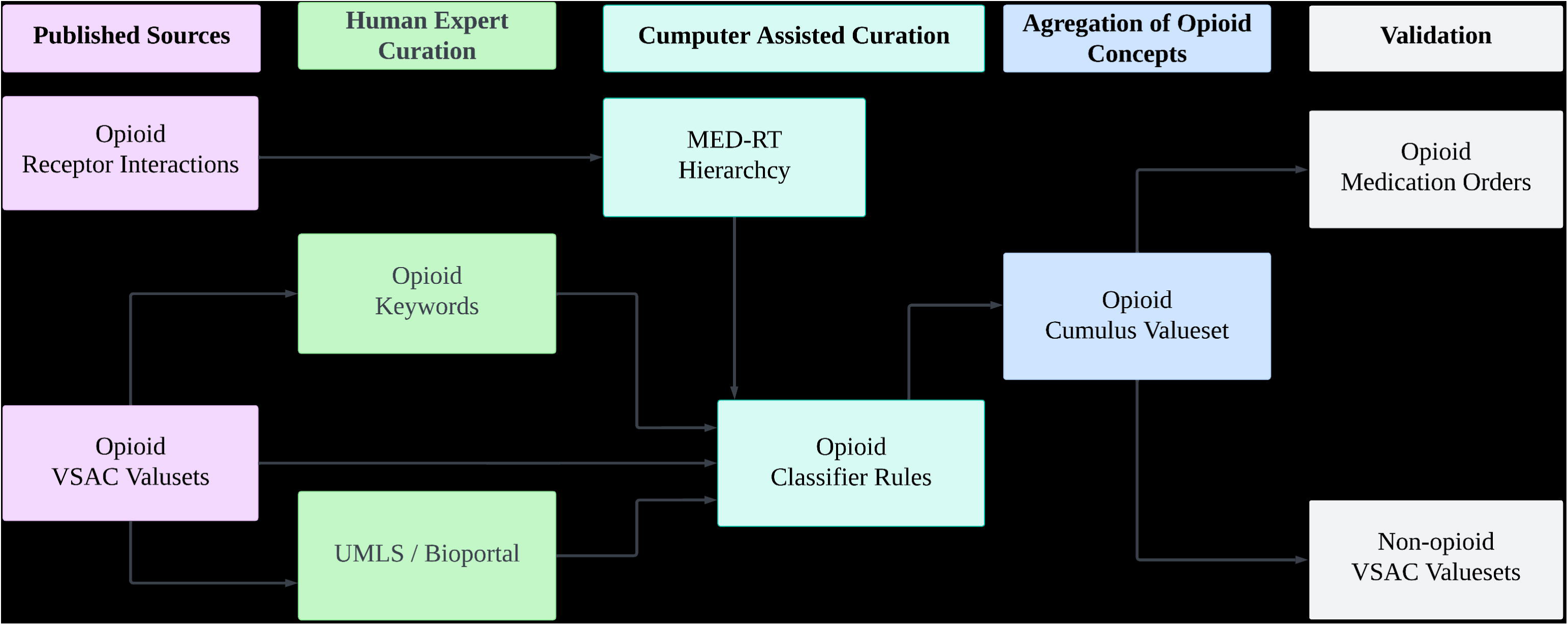
Methods to curate and validate the opioid Cumulus valueset. Published sources were human expert curated and expanded using computer assisted curation. The Cumulus opioid valueset contains the aggregation from these curated sources. Validation is measured against opioid medication orders and non-opioid VSAC valuesets. **Alt text:** Flowchart showing the process for curating and validating the Cumulus opioid valueset. Published sources, such as opioid receptor interactions and VSAC valuesets, undergo human expert curation and are expanded through computer-assisted curation using tools like MED-RT and BioPortal.

### Expert Manual Curation

The authors manually reviewed each opioid RxNorm concept from an opioid medications valueset[29] and verified each concept included an opioid ingredient in Bioportal. Each ingredient was then expanded using UMLS to include all RxNorm relationships[40] for all drug product formulations. In addition to ingredient-level curation, opioid keywords were curated to search for additional opioid medications. Common opioid-related terms, including both generic and brand names, were curated and refined iteratively to reduce false positives. The cumulus opioid keyword list was appended with a list of published VSAC valueset of opioid medication keywords[30].

### Computer Assisted Curation

Computer assisted curation was developed to expedite expert manual curation. Two computerized methods were developed: an opioid classifier and an MED-RT medication hierarchy of opioid receptor interactions. The opioid classifier rules were curated to select related opioid medications having similar opioid keywords, brand names, generic names, and multi-ingredient formulations by leveraging relationships such as “ingredient of” and “tradename of.” Additional rules were curated to disambiguate drug products with multiple ingredients such as cold medicines containing opioids. In total, the opioid classifier included keywords, ingredient-level relationships, and drug product formulations. The supplementary material contains the complete opioid classifier ruleset for each pair of RxNorm concepts, term types, and relationship attributes.

MED-RT was used to recursively include all RxNorm concepts in a hierarchy of opioid receptor interactions. MED-RT concepts were selected matching the keyword “opioid”. MED-RT provides a hierarchy of medication classes[32], namely mechanism of action (MoA), established pharmacologic class (EPC), and physiologic effects (PE). **Figure 2** illustrates the MED-RT partial hierarchy of Mu (μ) and Kappa (κ) opioid receptor interactions. Delta (δ) and nociceptin/orphanin FQ (NOP) receptors[41] are not included in the MED-RT hierarchy as there are currently no prescribable opioid medications for delta (δ) and NOP receptors.

**Figure 2:**
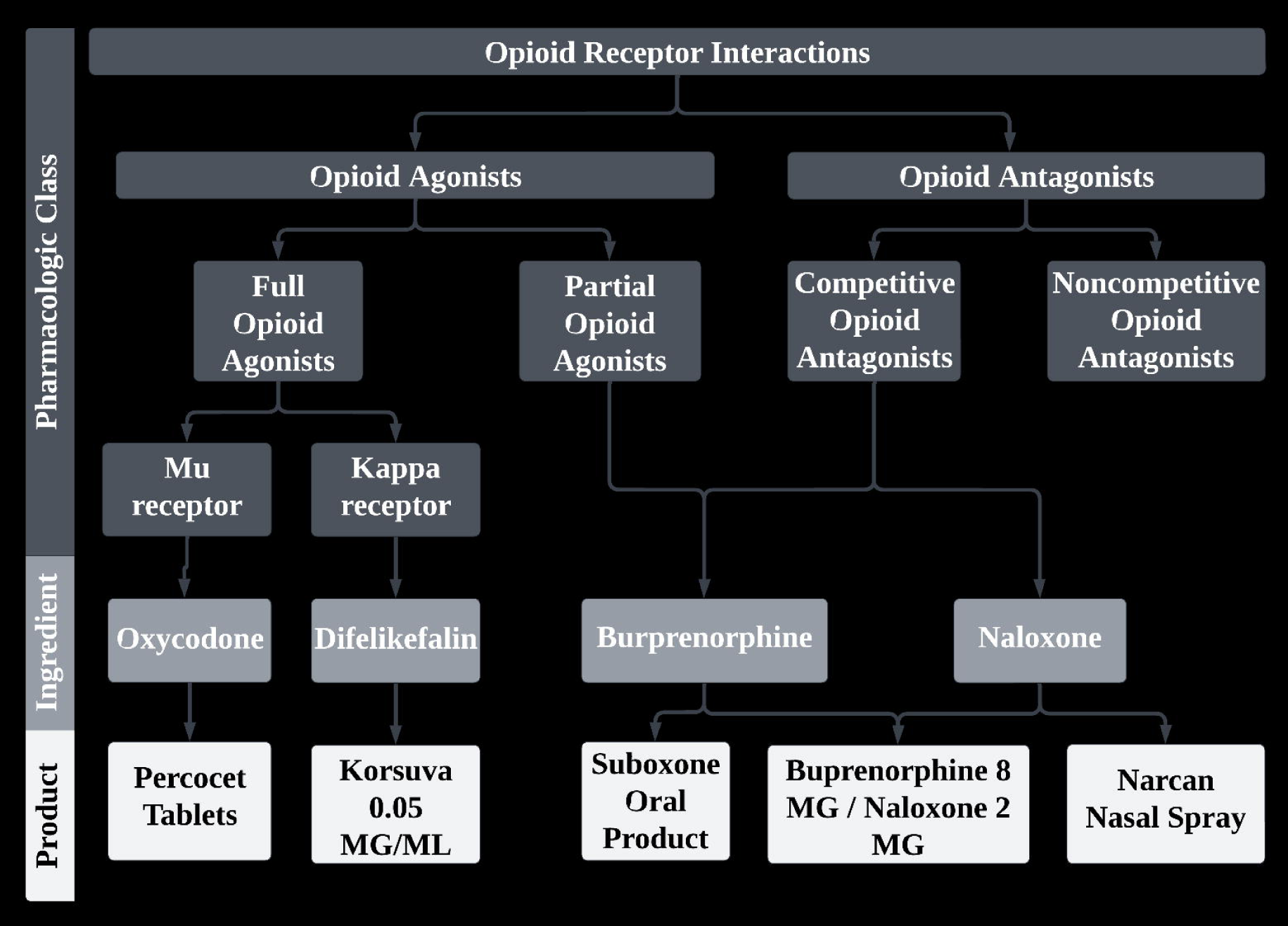
Example RxNorm medications in the MED-RT hierarchy of opioid receptor interactions. Opioid receptors hierarchically link RxNorm ingredients and RxNorm prescribable products. Shaded boxes denote MED-RT hierarchical concepts for the established pharmacologic class of opioid receptor interactions. Arrows denote direction of the hierarchical relationships. Unshaded boxes denote RxNorm medications. Bold denotes RxNorm ingredient concepts. Boxes on the bottom of the hierarchy provide examples of generic names, brand names, dose form, and strength. “Buprenorphine 8 MG / Naloxone 2 MG” illustrates how multiple ingredients can be combined in a single product. **Alt text:** Hierarchical diagram illustrating the MED-RT hierarchy for opioid receptor interactions. The diagram branches into opioid agonists and antagonists, which further divide into full, partial, competitive, and non-competitive subtypes. Examples of RxNorm medications include Oxycodone (a full opioid agonist for the Mu receptor) and Narcan (a competitive opioid antagonist). Additional medications such as Buprenorphine 8 MG / Naloxone 2 MG are shown as products with combined ingredients.

### Accuracy Measurements

Sensitivity and specificity were measured for the Cumulus opioid valueset against nine VSAC opioid valuesets[22–29,41] and three VSAC non-opioid valuesets[37–39]. **Figure 3** illustrates accuracy measurements. Sensitivity (recall) was measured as the percentage of VSAC opioid concepts recalled using the Cumulus opioid valueset, using formula true positive (TP) divided by the sum of TP plus false negative (FN) [i.e. TP / (TP + FN)]. Specificity (true negative rate) was measured using formula true negative (TN) divided by the sum of TN plus false positive (FP) [i.e. TN / (TN+FP)]. True positives were defined as opioid concepts in a VSAC opioid valueset. True negatives were defined as opioid concepts in a VSAC non-opioid valueset. False positives were defined as opioid concepts present in both the Cumulus opioid valueset and an VSAC non-opioid valueset. False negatives were defined as opioid concepts present in a VSAC opioid valueset that were not present in the Cumulus opioid valueset. Truth conflicts exist when the same RxNorm concepts is present in both a VSAC opioid and VSAC non-opioid valueset. VSAC truth conflicts were resolved by authors (MGW, AJM, PRVS) to confirm ground truth labeling.

**Figure 3:**
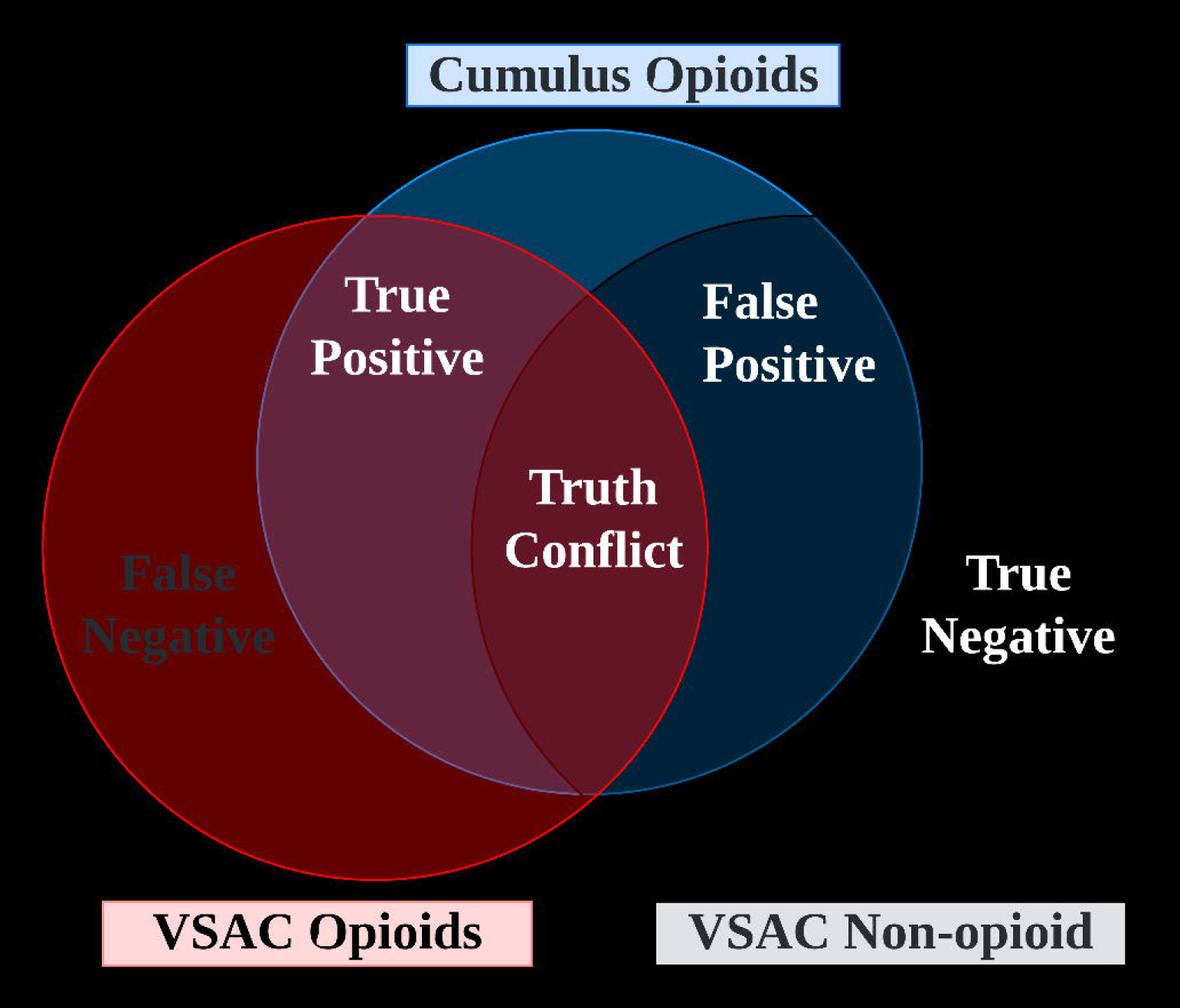
Accuracy of ground truth labels against the Cumulus and VSAC valuesets. True positives denote RXCUIs present in both Cumulus opioids and VSAC opioids. False negatives denote RXCUIs present only in VSAC opioids valuesets. False positive RXCUIs denote Cumulus opioids also present in VSAC non-opioids. True negative denotes RXCUIs present only in VSAC non-opioids. Truth conflict denotes RXCUIs present in both VSAC opioids and non-opioids. **Alt text:** Venn diagram illustrating the accuracy of Cumulus and VSAC valuesets. True positives are represented by the overlap between Cumulus opioids and VSAC opioids. False negatives refer to RXCUIs only present in VSAC opioids, while false positives denote RXCUIs present in Cumulus opioids but also found in VSAC non-opioids. True negatives represent RXCUIs only in VSAC non-opioids. A central section shows truth conflict, where RXCUIs exist in both VSAC opioids and non-opioids.

## RESULTS

Cumulus opioid valueset resulted in 8,926 unique opioid RxNorm concepts (**Figure 4**). To the authors’ knowledge, this is the largest publicly available opioid valueset conforming to standards federally mandated in every US-certified electronic health record system.

**Figure 4:**
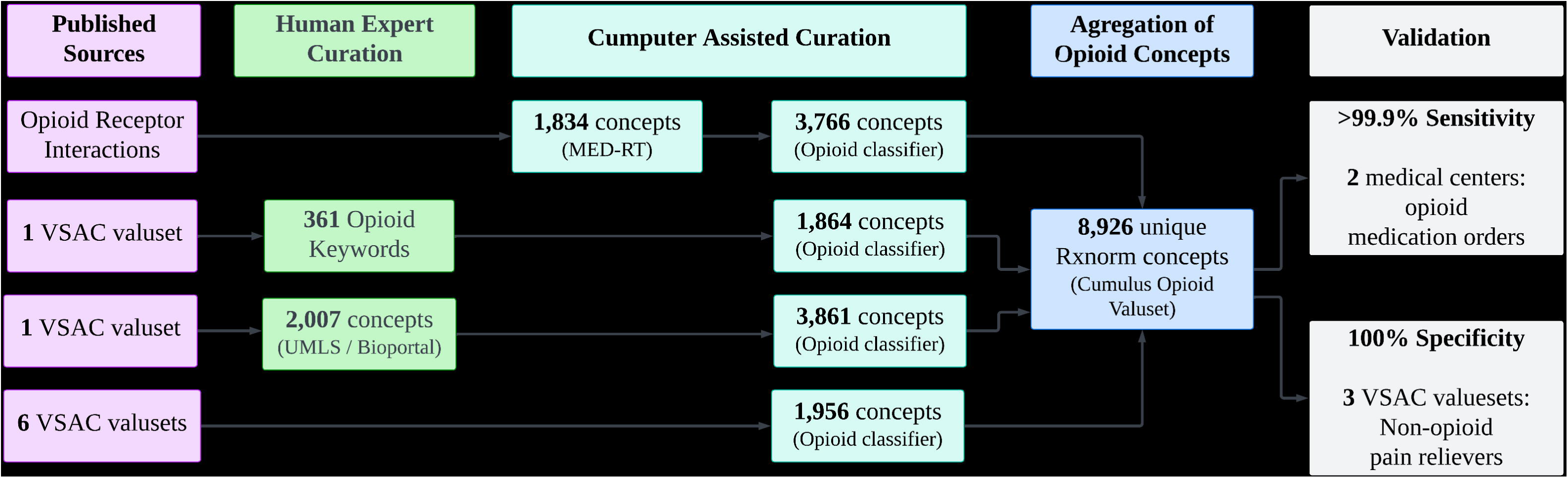
Cumulus opioid valueset curation and validation. Published sources of opioid receptor interactions, opioid keywords, and opioid medications are curated. Human expert curation included UMLS/BioPortal and curation of common opioid keywords. Cumulus assisted curation of opioid receptor interactions and published VSAC opioid valuesets. Cumulus opioid valueset resulted in 8,926 unique RxNorm concepts. Validation was performed against hospital opioid medication orders and valuesets of non-opioid pain-relievers. **Alt text:** Flowchart detailing the process of curating and validating the Cumulus opioid valueset. Human expert curation incorporates opioid receptor interactions, keywords, and concepts from UMLS/BioPortal. The process expands through computer-assisted curation, adding concepts from the MED-RT hierarchy and various VSAC valuesets. The final aggregation yields 8,926 unique RxNorm concepts, validated against opioid medication orders (99.9% sensitivity) and non-opioid valuesets (100% specificity).

Manual curation of 349 opioid medications[29] using UMLS/BioPortal yielded an expanded list of 2,007 RxNorm concepts. Opioid classifier rules were then used to select an additional set of 3,861 related concepts. MED-RT hierarchy included 1,834 concepts with a known opioid receptor interaction[35], which were then related to an additional 3,766 concepts using the opioid classifier rules. Opioid keywords were used to select 1,864 concepts from published VSAC opioid valuesets, which were then expanded to include 1,520 additional concepts. In total, 8,926 unique RxNorm concepts were selected using manual curation, opioid receptor interactions, opioid keywords, and opioid classifier rules of related RxNorm concepts.

### Validation

Cumulus opioid valueset was tested against multiple opioid and non-opioid valuesets, yielding >99.9% sensitivity and 100% specificity. Cumulus recalled 1,193 of 1,994 unique RxNorm concepts. Of these, Cumulus recalled all 228 concepts for opioid oral tablets and patches from Brigham and Women’s Hospital[42]. Cumulus recalled 1007 of 1137 concepts for opioids in the UC Davis Health set of opioid prescriptions. Upon manual review, only 1 of the 130 concepts missing opioid concepts was determined to be a prescribable opioid medication. The single truth conflict was due to the UC Davis Epic EHR medications grouper inadvertently including individual ingredients such as aspirin, ibuprofen, and guaifenesin that are commonly prescribed with codeine.

Cumulus opioid valueset specificity was 100% compared to three published VSAC valuesets for non-opioids[37–39]. None of the 3877 non-opioid RXCUIs were present in the Cumulus opioid valueset. Additional validation was performed against 2048 non-opioid concepts in a MED-RT hierarchy containing RxNorm concepts for classes: NSAID, barbiturate, and/or benzodiazepine. Confirmed by manual review, the Cumulus opioid valueset contained zero false positives from any non-opioid valueset[22].

VSAC validation rules were applied to the set of 8,926 RxNorm concepts resulting in 4,123 active concepts and 4,803 inactive concepts. Both active and inactive concepts are provided to readers to enable historical analysis of opioid prescriptions since the start of the opioid crisis.

## DISCUSSION

To the authors’ knowledge, this study presents the largest publicly available valueset of opioid medications using the RxNorm standard. The intended use of the Cumulus opioid valueset is to quickly measure opioid population health statistics across multiple healthcare organizations in parallel[9]. Cumulus is designed for use with US core standards for interoperability(USCDI) [43] with over 100 data elements[44] relevant to opioid studies, especially medications[45], conditions[46,47], vital signs[48], laboratory results[49], diagnostic reports[50], observations[51], clinical notes[52], and demographics[53] across a wide range of encounter settings[54]. When used with the USCDI data, Cumulus opioid valueset may be used to measure frequency of opioid prescriptions[55], trajectories of opioid use[56], treatment of opioid use disorders[57,58], and other opioid related population health statistics.

There were several limitations in this study. The Cumulus opioid valueset includes RxNorm opioid medication prescribed by a healthcare professional. Opioids acquired outside[59] of the healthcare system cannot be measured using prescriptions alone[60]. It was beyond the scope of this study to evaluate case definitions of substance use disorders using diagnosis concepts[61] and laboratory results[62]. Despite best efforts, it should be expected that not all cases of opioid use will be counted. The unregulated opioid supply frequently changes ingredient mixtures and chemical structures thus limiting detection using standardized toxicology screens. To estimate the true opioid use prevalence from the apparent opioid use prevalence[63], clinical notes[52] could be reviewed for a small sample of the population to adjust prevalence estimates.

## DATA SHARING

Data and code are freely available under the Apache-2.0 license. Code for building the Cumulus opioid valueset is available at https://github.com/smart-on-fhir/cumulus-library-opioid-valueset. Supplementary data is available at https://github.com/smart-on-fhir/cumulus-library-opioid-valueset-supplement.

## ACKNOWLEDGMENTS

Authors thank the UC Davis pharmacy and informatics teams with special thanks to Eugene Kang and Momeema Ali.

