## Supplemental File 1 for "Development and validation of a near-comprehensive RxNorm valueset of opioid medications"

### **Valueset References**

| **Count** | **Steward** | **Abbreviation** | **Contains** | **ValueSet Identifier** |
| --- | --- | --- | --- | --- |
| 1612 | CancerLinQ | CLQ | Opioids | 2.16.840.1.113762.1.4.1116.449 |
| 1004 | IMPAQ International | Impaq | All opioids | 2.16.840.1.113762.1.4.1196.87 |
| 836 | American College of Emergency Physicians / AMA-PCPI | ACEP | Opioid meds | 2.16.840.1.113762.1.4.1106.68 |
| 753 | MITRE | MITRE | Opioid pain medications | 2.16.840.1.113762.1.4.1032.34 |
| 455 | Emergency Care Research Institute | ECRI | All prescribable opioids used for pain control including Inactive Medications | 1.3.6.1.4.1.6997.4.1.2.234.999.3.2 |
| 379 | Lantana | Lantana | Schedule II, III and IV opioid medications | 2.16.840.1.113762.1.4.1046.241 |
| 349 | Mathematica | Math | Opioid medications | 2.16.840.1.113883.3.3157.1004.26 |
| 228 | Brigham and Women’s Hospital | BWH | Opioid (oral tablets and patches only) | 2.16.840.1.113762.1.4.1206.12 |
| 38 | CliniWiz | CliniWiz | Opioid medication Keywords | 2.16.840.1.113762.1.4.1200.163 |

**Table S1: Opioid valuesets.** Count is the number of RxNorm unique concept identifiers. Each opioid valueset is named by the steward who curated and published the list of RxNorm codes. Abbreviation is the short name used in supplementary tables.

| **Count** | **Steward** | **Abbreviation** | **Contains** | **ValueSet Identifier** |
| --- | --- | --- | --- | --- |
| 1968 | MITRE | non-MITRE | Non opioid from ATC | 2.16.840.1.113762.1.4.1032.291 |
| 1906 | MD Partners | non-MD | Non opioid pain medications | 2.16.840.1.113762.1.4.1021.73 |
| 1305 | CancerLinQ | non-CLQ | Non opioid analgesics | 2.16.840.1.113762.1.4.1260.173 |

**Table S2: Non opioid valuesets.** Count is the number of RxNorm unique concept identifiers. Each non opioid valueset is named by the steward who published the list of RxNorm codes. Abbreviation is the short name used in supplementary tables. The prefix “non” denotes “non opioid” valueset.

| **Count** | **Steward** | **Abbreviation** | **Contains** | **Description** |
| --- | --- | --- | --- | --- |
| 9223 | Cumulus | Opioid | Opioids | Cumulus Opioid test valueset for accuracy measurement prior to final human expert review. |
| 2048 | Cumulus | non-MEDRT | Non-opioids | MED-RT classes: nonsteroidal anti-inflammatory drug (NSAID), barbiturate, and/or benzodiazepine. |
| 2007 | Cumulus | Bioportal | Opioids | Manually curated list of opioids developed by looking up known opioid ingredients in the NCBO bioportal online resource. |
| 1850 | Cumulus | MED-RT | Opioids | MED-RT drugs with known "Opioid Receptor Interactions" mechanism of action (MoA) or pharmacologic class (EPC). |
| 1136 | Cumulus | UCDavis | Opioids | Opioid medication orders in Epic EHR from June 2023 to June 2024. |

**Table S3: Cumulus developed valuesets.** Count is the number of RxNorm unique concept identifiers. Each non opioid valueset is named by the steward who published the list of RxNorm codes. Abbreviation is the short name used in supplementary tables.
