## Supplemental File 2 for "Development and validation of a near-comprehensive RxNorm valueset of opioid medications"

### **UMLS/BioPortal Curation**

Each of the 349 RxNorm codes in the Mathematica[[1]](https://paperpile.com/c/8AJRF1/d0R6) “Opioid medications” valueset were manually reviewed at the ingredient level. From the 349 RxNorm codes, 167 unique ingredients were curated, which in turn were related to 7,703 RxNorm codes having relationship attribute “ingredient of”. Deduplication resulted in 2,007 unique RxNorm codes. Compared to the CancerLinQ[[2]](https://paperpile.com/c/8AJRF1/hmNb), the largest existing opioid value set (1,612 codes), the manually curated value set using Bioportal was larger (2,007 codes). This manually curated value set included larger numbers of matched RxNorm codes in every evaluated VSAC opioid value set.

### **Opioid Keywords Curation** Keywords matches are available at <https://github.com/smart-on-fhir/cumulus-library-opioid-valueset>.

The process of keyword curation began with a manual examination of commonly used terms, including brand names associated with opioid-use medications, derived from the curated Bioportal list. Initially, the keyword list encompassed basic names of brand-name opioids (e.g., "Atuss"), aimed at capturing various versions of the brand (e.g., Atuss EX, Atuss HD, Atuss MS). This preliminary approach was intended to ensure comprehensive identification of all iterations of specific opioid medications.

During the preliminary testing phase of the keyword curation, it became apparent that the general brand names for cold-cough opioid medications required greater specificity. The initial set of keywords predominantly flagged cold-cough medications that did not contain opioids, thus leading to a high rate of false positives. This observation necessitated a refinement of our keyword strategy.

Through an iterative refinement process, we identified several keywords that were generating false positives. Specifically, terms such as "opium," "vidone," and "bitex," despite their association with opioids, were frequently part of non-opioid medication names. For instance, the keyword "opium" flagged medications such as "ipratropium" and "Tropium," neither of which are opioid medications. This issue highlighted the need for more precise keywords to enhance the accuracy of opioid medication identification.

To address these inaccuracies, we adopted a two-fold approach. For keywords that could be specified further, such as "vidone," we included additional descriptors to narrow down the context (e.g., "vidone oral" and "vidone pill"). Conversely, for keywords that could not be sufficiently refined without compromising accuracy, such as "opium," we opted to remove them entirely from the list.

This refined approach to keyword curation, through rigorous testing and adjustment, ensured that our keyword list was both comprehensive and precise, effectively minimizing false positives while maintaining the ability to identify a wide range of opioid-use medications. The final curated keyword list thus represents a robust tool for accurately flagging opioid-related terms within the context of medication identification.

| **Keyword Length** | **Keyword count** | **RxNorm codes** | **String labels** |
| --- | --- | --- | --- |
| 0 (No match) | 0 | 380560 | 843159 |
| 4 | 1 | 3 | 3 |
| 5 | 20 | 181 | 343 |
| 6 | 48 | 564 | 1048 |
| 7 | 75 | 3207 | 6880 |
| 8 | 79 | 4666 | 12329 |
| 9 | 55 | 898 | 2275 |
| 10 | 45 | 647 | 1514 |
| 11 | 24 | 219 | 382 |
| 12 | 7 | 32 | 45 |
| 13 | 3 | 253 | 791 |
| 14 | 2 | 10 | 18 |
| 15 | 2 | 11 | 18 |

**Table: Keyword Length Distribution**. In total, 361 opioid keyword phases were manually curated. Keyword length is the number of characters. Keyword count is the number of keywords of the specified keyword length. RxNorm code column denotes number of unique concept identifiers (RXCUI). String labels refers to the number of unique RxNorm supplied labels, which may be ingredient, brand name, or other drug product description.

### **Opioid Classifier Rules Curation**

Classifier rules were developed to include brand names, generic names, prescribable names, and other formulations, as well as medications with exact ingredient relationships. The selection and formulation of the rules were performed iteratively to minimize false positives and maintain relevance to opioid medications. Classifier rules use previously curated RxNorm Relationship Attributes[[3]](https://paperpile.com/c/8AJRF1/vNGq) (RELA).

Specific relationships (RELA) and term types (TTY) were provided in the RxNorm database release version July 01, 2024 . For brand names (BN), we included the relationship "BN reformulated_to BN" because it maintains consistency without broadening or narrowing the scope of classification. This was exemplified by the relationship 'Midol PM' reformulated_to 'Midol PM Reformulated Apr 2011'. Similarly, "BN reformulation_of BN" was included to ensure that reformulated versions were accurately captured. The relationship "BN tradename_of IN" was included based on keywords, recognizing that while this inclusion is generally reliable, it could become ambiguous with repeated iterations. An example is 'Tylenol' tradename_of 'Acetaminophen'. We also included "BN has_precise_ingredient PIN" based on keywords, anticipating that precise ingredients were already captured in our keyword list, as shown by 'Tylenol PM' has_precise_ingredient 'Diphenhydramine Hydrochloride'. Finally, relationships like "BN ingredient_of SBD/SBDC/SBDF/SBDG" were included to ensure the classification of brand-specific doses, exemplified by 'Tylenol' ingredient_of 'Acetaminophen 325 MG Oral Tablet [Tylenol]'.

For dose form (DF) and dose form group (DFG) relationships, we decided not to include them because they do not provide specific information about the medication’s opioid content. In contrast, for pack (BPCK) and group pack (GPCK) relationships, we included "BPCK has_tradename GPCK" and "BPCK contains SBD/SCD" to ensure the inclusion of related pack names and specific dose forms within packs. The relationship "GPCK has_tradename BPCK" was also included to maintain consistency with pack naming conventions, while "GPCK contains SCD" was included based on keywords due to the potential for including non-opioid medications.

In terms of ingredient (IN) and precise ingredient (PIN) relationships, we included "IN has_tradename BN" based on keywords to ensure proper association with brand names, exemplified by 'Acetaminophen' has_tradename 'Tylenol'. The relationship "IN part_of MIN" was included based on keywords to reflect complex ingredient relationships, such as 'Acetaminophen' part_of 'Acetaminophen/Diphenhydramine'. We also included "IN has_form PIN" and "IN ingredient_of SCDC/SCDF/SCDFP/SCDG" based on keywords to maintain ingredient specificity. For multi-ingredient (MIN) relationships, "MIN has_part IN/PIN" was included based on keywords, recognizing the potential to yield non-opioids and necessitating keyword verification. An example is 'Acetaminophen/Diphenhydramine' has_part 'Acetaminophen'. The relationship "MIN ingredients_of SCD" was included to ensure the inclusion of combined dose forms.

For specific dose form (SBD) relationships, we included "SBD has_ingredient BN" to ensure association with brand names. We also included "SBD contained_in BPCK/GPCK" to ensure the inclusion of dose forms within packs. Relationships such as "SBD quantified_form_of SBD" and "SBD consists_of SBDC" were included to maintain consistency within quantified forms and ensure the inclusion of dose form components. The relationships "SBD isa SBDF/SBDFP/SBDG" were included based on keywords to prevent false positives with multi-word brand names, and "SBD tradename_of SCD" was included to ensure proper association with dose forms.

For specific clinical dose form (SCDC), specific dose form (SCDF), and specific dose form pack (SCDFP) relationships, we included "SCDC has_ingredient IN/PIN" based on keywords to ensure accurate ingredient inclusion. We also included "SCDC constitutes SBD/SCD" and "SCDC has_tradename SBDC" to ensure proper association with brand doses and specific clinical doses.

To ensure robustness and prevent errors, we implemented safety nets to exclude dose forms (DF) and dose form groups (DFG) as TTY. This logic helps prevent errors and maintains the classifier's accuracy.

### **Opioid Classifier Rules List**

The Opioid classifier implements rules for expanding from a known opioid (concept 1) to a related candidate ( concept 2). Both the known opioid concept and related concept have defined RxNorm Term Types[[4]](https://paperpile.com/c/8AJRF1/FXU2). The pair are related by the RxNorm Relationship Attribute[[3]](https://paperpile.com/c/8AJRF1/vNGq) (RELA).

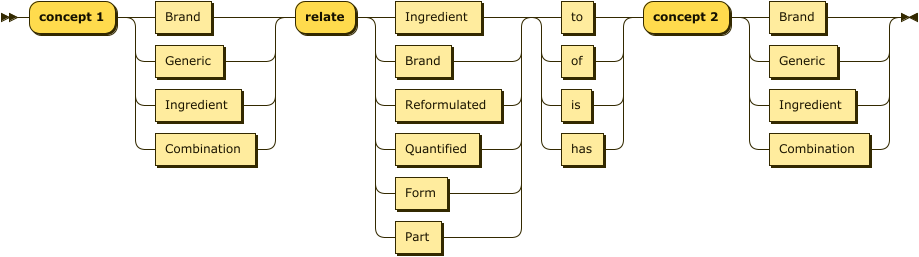

**Figure: Opioid Related Concepts**. Shown: generalization of relationship attributes (RELA) and concept term types (TTY) used to define Opioid classifier rules.

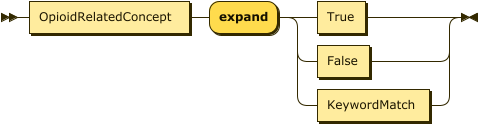

**Figure: Opioid Classifier Rule.** Each opioid classifier rule determines if related concept 2 should be included in the list of opioids. Concept 1 is already assumed to be an opioid. True: include concept 2 in the opioid valueset. False: do not include concept 2 in the opioid valueset. KeywordMatch: include concept 2 if and only if there the concept matches an Opioid keyword.

| **# TTY1** | **RELA** | **TTY2** | **INCLUDE** |
| --- | --- | --- | --- |
| **BN** | reformulated_to | BN | TRUE |
| **BN** | reformulation_of | BN | TRUE |
| **BN** | tradename_of | IN | Keyword |
| **BN** | has_precise_ingredient | PIN | Keyword |
| **BN** | ingredient_of | SBD | TRUE |
| **BN** | ingredient_of | SBDC | TRUE |
| **BN** | ingredient_of | SBDF | TRUE |
| **BN** | ingredient_of | SBDG | TRUE |
| **BPCK** | has_dose_form | DF | FALSE |
| **BPCK** | tradename_of | GPCK | TRUE |
| **BPCK** | contains | SBD | TRUE |
| **BPCK** | contains | SCD | TRUE |
| **DF** |  |  | FALSE |
| **DFG** |  |  | FALSE |
| **GPCK** | has_tradename | BPCK | TRUE |
| **GPCK** | has_dose_form | DF | FALSE |
| **GPCK** | contains | SCD | Keyword |
| **IN** | has_tradename | BN | Keyword |
| **IN** | part_of | MIN | Keyword |
| **IN** | has_form | PIN | Keyword |
| **IN** | ingredient_of | SCDC | Keyword |
| **IN** | ingredient_of | SCDF | Keyword |
| **IN** | ingredient_of | SCDG | Keyword |
| **IN** | boss_of | SCDFP | Keyword |
| **MIN** | has_part | IN | Keyword |
| **MIN** | has_part | PIN | Keyword |
| **MIN** | ingredients_of | SCD | TRUE |
| **PIN** | precise_ingredient_of | BN | TRUE |
| **PIN** | form_of | IN | Keyword |
| **PIN** | part_of | MIN | TRUE |
| **PIN** | precise_ingredient_of | SCDC | TRUE |
| **PIN** | boss_of | SCDFP | TRUE |
| **SBD** | has_ingredient | BN | TRUE |
| **SBD** | contained_in | BPCK | TRUE |
| **SBD** | has_dose_form | DF | FALSE |
| **SBD** | quantified_form_of | SBD | TRUE |
| **SBD** | has_quantified_form | SBD | TRUE |
| **SBD** | consists_of | SBDC | TRUE |
| **SBD** | isa | SBDF | Keyword |
| **SBD** | isa | SBDFP | Keyword |
| **SBD** | isa | SBDG | Keyword |
| **SBD** | tradename_of | SCD | TRUE |
| **SBD** | consists_of | SCDC | TRUE |
| **SBDC** | has_ingredient | BN | TRUE |
| **SBDC** | constitutes | SBD | TRUE |
| **SBDC** | tradename_of | SCDC | TRUE |
| **SBDF** | has_ingredient | BN | TRUE |
| **SBDF** | has_dose_form | DF | FALSE |
| **SBDF** | inverse_isa | SBD | TRUE |
| **SBDF** | isa | SBDG | TRUE |
| **SBDF** | tradename_of | SCDF | TRUE |
| **SBDF** | has_form | SBDFP | TRUE |
| **SBDFP** | form_of | SBDF | TRUE |
| **SBDFP** | tradename_of | SCDFP | TRUE |
| **SBDFP** | inverse_isa | SBD | TRUE |
| **SBDG** | has_ingredient | BN | TRUE |
| **SBDG** | has_doseformgroup | DFG | FALSE |
| **SBDG** | inverse_isa | SBD | TRUE |
| **SBDG** | inverse_isa | SBDF | TRUE |
| **SBDG** | tradename_of | SCDG | Keyword |
| **SBDG** | tradename_of | SCDGP | TRUE |
| **SCD** | contained_in | BPCK | TRUE |
| **SCD** | has_dose_form | DF | FALSE |
| **SCD** | contained_in | GPCK | TRUE |
| **SCD** | has_ingredients | MIN | TRUE |
| **SCD** | has_tradename | SBD | TRUE |
| **SCD** | quantified_form_of | SCD | TRUE |
| **SCD** | has_quantified_form | SCD | TRUE |
| **SCD** | consists_of | SCDC | TRUE |
| **SCD** | isa | SCDF | Keyword |
| **SCD** | isa | SCDG | Keyword |
| **SCD** | isa | SCDFP | TRUE |
| **SCDC** | has_ingredient | IN | Keyword |
| **SCDC** | has_precise_ingredient | PIN | Keyword |
| **SCDC** | constitutes | SBD | TRUE |
| **SCDC** | has_tradename | SBDC | TRUE |
| **SCDC** | constitutes | SCD | TRUE |
| **SCDF** | has_dose_form | DF | FALSE |
| **SCDF** | has_ingredient | IN | Keyword |
| **SCDF** | has_tradename | SBDF | Keyword |
| **SCDF** | inverse_isa | SCD | TRUE |
| **SCDF** | isa | SCDG | Keyword |
| **SCDF** | has_form | SCDFP | TRUE |
| **SCDFP** | inverse_isa | SCD | TRUE |
| **SCDFP** | has_tradename | SBDFP | TRUE |
| **SCDFP** | form_of | SCDF | TRUE |
| **SCDFP** | isa | SCDGP | TRUE |
| **SCDFP** | has_boss | IN | Keyword |
| **SCDFP** | has_boss | PIN | Keyword |
| **SCDG** | has_doseformgroup | DFG | FALSE |
| **SCDG** | has_ingredient | IN | Keyword |
| **SCDG** | has_tradename | SBDG | Keyword |
| **SCDG** | inverse_isa | SCD | TRUE |
| **SCDG** | inverse_isa | SCDF | TRUE |
| **SCDGP** | has_tradename | SBDG | Keyword |
| **SCDGP** | inverse_isa | SCDFP | TRUE |
| **SCDGP** | form_of | SCDG | Keyword |

**Table: Opioid Classifier Rule List.** TTY1 and TTY2 denote RxNorm term types[[4]](https://paperpile.com/c/8AJRF1/FXU2) for a pair concept 1 and concept 2, respectively. Concept 1 is assumed to contain an opioid. Concept 2 opioid classification is unknown. RELA denotes the RxNorm relationship[[3]](https://paperpile.com/c/8AJRF1/vNGq) between RxNorm concept pairs. Expand denotes if the concept pair should be expanded. TRUE classifies concept 2 as an opioid. FALSE classifies concept 2 as not an opioid. Keyword applies a keyword matching rule: classify concept 2 as an opioid only if concept 2 matches a known opioid keyword. If no match is found, concept 2 is classified as not an opioid.

3	Appendix 1 - RxNorm Relationships (RELA). Published Online First: 1 September 2023.

4	Appendix 5 - RxNorm Term Types (TTY). Published Online First: 1 September 2023.
